# Use of Ketamine Infusions for Treatment of Complex Regional Pain Syndrome: A Systematic Review

**DOI:** 10.1101/2021.04.20.21255840

**Authors:** Anand S. Patil, Ahish Chitneni, Suhani Dalal, Joe Ghorayeb, Yolanda Pham, Gregory Grigoropoulos

**Affiliations:** Touro University California, College of Osteopathic Medicine; A.T Still University School of Osteopathic Medicine in Arizona (SOMA); University of Medicine and Health Sciences; Loyola University Chicago, Stritch School of Medicine

## Abstract

**Background:** This systematic review aims to review clinical studies on the use of ketamine infusion for patients with treatment-resistant Complex Regional Pain Syndrome (CRPS).

**Methods:** The following systematic review was registered on PROSPERO (CRD42021228470). Studies for the systematic review were identified through 3 databases; PubMed, CINAHL, and Cochrane Review. Inclusion criteria for studies consisted of randomized clinical trials or cohort studies that conducted trials on the use of ketamine infusion for pain relief in patients with Complex Regional Pain Syndrome (CRPS). Exclusion criteria for studies included any studies that were systematic review, meta-analyses, case reports, literature reviews, or animal studies. In the included studies, the primary outcome of interest was post drug administration pain score.

**Results:** In this systematic review, 14 studies met the inclusion criteria and were reviewed. In these studies, the dosage of ketamine infusion used ranged from 0.15 mg/kg to 7mg/kg with primary indication being treatment of Complex Regional Pain Syndrome (CRPS). In 13 of the studies, ketamine infusion resulted in a decrease in pain scores and relief of symptoms.

**Conclusions:** Patients who received Ketamine infusion for treatment-resistant CRPS self-reported adequate pain relief with treatment. This suggests that ketamine infusion may be a useful form of treatment for patients with no significant pain relief with other conservative measures. Future large-scale studies, including randomized double-blind placebo-controlled trials on the use of ketamine infusion for CRPS must be conducted in a large-scale population to further assess the effectiveness of ketamine infusion in these populations.

## Introduction

Complex Regional Pain Syndrome (CRPS) presents with pain in the extremities following trauma. The typical patient is a middle aged female, with upper limb insult causing lasting pain and/or concurrent dysfunction in the sensory, motor, autonomic, and trophic domains.^1^ CRPS is further classified into type I and II, the latter involving identification of a specific nerve lesion.^2^ CRPS makes up approximately 1.2% of diagnosis of chronic pain within the American patient population and 7% of patients post trauma.^3^ From a financial perspective, CRPS patients on average spent more on pain medications after their diagnosis. Due to its ill-defined characteristics, CRPS often requires management by a multidisciplinary team thus faring more expensive than treatment of other chronic pain diagnoses. Compared to baseline before diagnosis, patients have a 2.17-fold increase in prescription cost and 2.56-fold increase in total cost annually ranging from $3888 to $4845.^4^ Furthermore, only approximately 40% of patients diagnosed with CRPS will return to full-time work, proving to have an extensive impact on socioeconomic factors.^5^

Among the diverse therapies available for CRPS including immunomodulation, hyperbaric oxygen therapy and psychological therapy - medicinal approaches have yielded the most efficacy. In particular, the open-channel NMDA blocker ketamine, with its greater safety profile than other interventions, is beneficial when standard treatments have failed.^6^ Ketamine is a phencyclidine (PCP) derivative that initially became commercially available for human use in 1970 as a rapid-acting intravenous (IV) anesthetic,^7^ and is currently classified by the Food and Drug Administration as an anesthetic induction agent in doses ranging from 1 to 4.5 mg/kg.^8^ Ketamine has proven to be a desirable drug, despite its induction of dissociative effects^9^ and abuse potential.^10^ It is favorable due to its short half-life and lack of clinically significant respiratory depression.^11^ In addition to its anesthetic effects, ketamine possesses analgesic,^12^ anti-inflammatory,^13^ and antidepressant activity.^14^ These characteristics bode well for the treatment of CRPS for which treatment is approached through a biopsychosocial model of chronic pain.^15^

### Pharmacology & Mechanism of Action of Ketamine

Ketamine is composed of a chiral center at the C-2 carbon of the cyclohexanone ring, so that two enantiomers exist: *S*(+)-ketamine and *R*(-)-ketamine. The *S*-enantiomer has greater pharmacological potency due to greater affinity for the PCP binding site on the *N*-methyl-d-aspartate (NMDA) receptor.^16^ Accordingly, preparations containing only the S-enantiomer are desirable.

Traditionally, ketamine is administered via IV or intramuscular routes. Alternative routes of administration include insufflation/intranasal, inhalational, oral, topical, epidural, and rectal. It is both water and lipid soluble. These qualities allow for its rapid distribution throughout the body and ability to cross the blood-brain barrier. The predominant route of metabolism is via hepatic cytochrome P450 enzymes, with 12% remaining protein bound in plasma.^16^ Ketamine’s half-life in plasma is approximately 2.5 to 3 hours, rapidly metabolizing to norketamine, hydroxynorketamine and dehydronorketamine. Approximately 4% of unmetabolized drug is excreted via urine.^16^

The therapeutic range of ketamine makes it one of the safest sedative agents for most emergency clinical and preclinical situations. In low doses, it causes analgesia and sedation and in high doses, it produces general anesthesia. Administration of ketamine typically increases heart rate, systolic and diastolic blood pressure, salivary and tracheobronchial secretions, and bronchodilation.^17^ It has minimal effects on airway reflexes and respiratory rate. Currently, three pain societies recommend intravenous dosing of ketamine for chronic pain at 0.5 to 2 mg/kg for 1-day outpatient or 3- to 5-day inpatient treatment with higher doses titrated to effect. ^18^

Ketamine exerts its effects through a variety of pathways. Its primary mechanism is as a non-competitive antagonist at the PCP binding site of the NMDA receptors in the central nervous system (CNS).^19^ At this site, it decreases the frequency of channel opening and duration of time spent in the active, open state.^20^ The NMDA receptor is a ligand-gated channel whose major endogenous agonist is glutamate, the predominant excitatory neurotransmitter in the CNS. Inhibition of the NMDA receptor results in decreased neuronal activity.

Ketamine also acts on other non-NMDA pathways that affect pain and mood regulation. It acts as an antagonist of nicotinic and muscarinic cholinergic receptors as well as blocking sodium and potassium channels. Ketamine activates high-affinity D_2_ dopamine receptors and l-type voltage-gated calcium channels, promotes γ-aminobutyric acid A (GABA-A) signaling, and enhances descending modulatory pathways.^16,21,22^

Given the high co-prevalence rate of chronic pain and depression, the antidepressant effects of ketamine have generated great interest in the psychiatric community. The mood-enhancing effects of ketamine have been shown to emerge about 4 hours after intravenous administration, well after the drug has been cleared from the bloodstream. This suggests a neuromodulatory effect that persists for 1 to 2 weeks.^23^

## Methods

The following systematic review was registered on the PROSPERO website (CRD42021228470) and the systematic review follows the guidelines listed by the Preferred Reporting Items for Systematic Reviews and Meta-Analyses (PRSIMA) statement. In the review, studies describing the use of ketamine infusion for Complex Regional Pain Syndrome were researched to validate the use of the treatment.

### Search Strategy

An electronic literature search using three databases was conducted by the team. The databases PubMed, Cochrane Review, and CINAHL were searched using the terms “Ketamine” OR “Ketamine infusion” AND “CRPS” OR “Complex Regional Pain Syndrome” to find peer-reviewed clinical studies. No date limiters were used. Initial search resulted in 101 non duplicate entries from the three databases. After application of inclusion/exclusion, 87 articles were excluded from the search and 14 studies were included in the study.

### Inclusion & Exclusion Criteria

Inclusion criteria consisted of all studies that were randomized controlled trials or studies that observed the effect of the use of ketamine infusion in pain relief for patients with complex regional pain syndrome (CRPS). Exclusion criteria consisted of nonpeer-reviewed studies, systematic or meta-analysis reviews, case reports, and animal studies. All articles were reviewed by both reviewer AP and reviewer AC separately and any disagreements were resolved by tiebreaker reviewer SD. After the review process, both reviewer AP and AC had a 95% agreement rate for the studies to be included.

## Data Extraction

Of the studies to be included in the paper, the following data was extracted for discussion: 1) length of ketamine infusion, 2) follow up period, 3) dose of ketamine used, 4) outcomes of interest, and 5) post-procedural pain scores.

## Results

### Study Characteristics

A total of 101 articles were identified as meeting the keyword search between the databases. After removing duplicates, 84 articles were screened, and 56 did not meet inclusion criteria based on their respective titles and abstracts. The remaining 28 articles were reviewed in full-text and an additional 14 articles were excluded due to meeting exclusion criteria. Fourteen total articles were selected to be included in this review (Figure 1). A total of 455 patients were analyzed between the 14 included studies. Sample sizes ranged from 4 to 114 human subjects.

**FIGURE 1.**
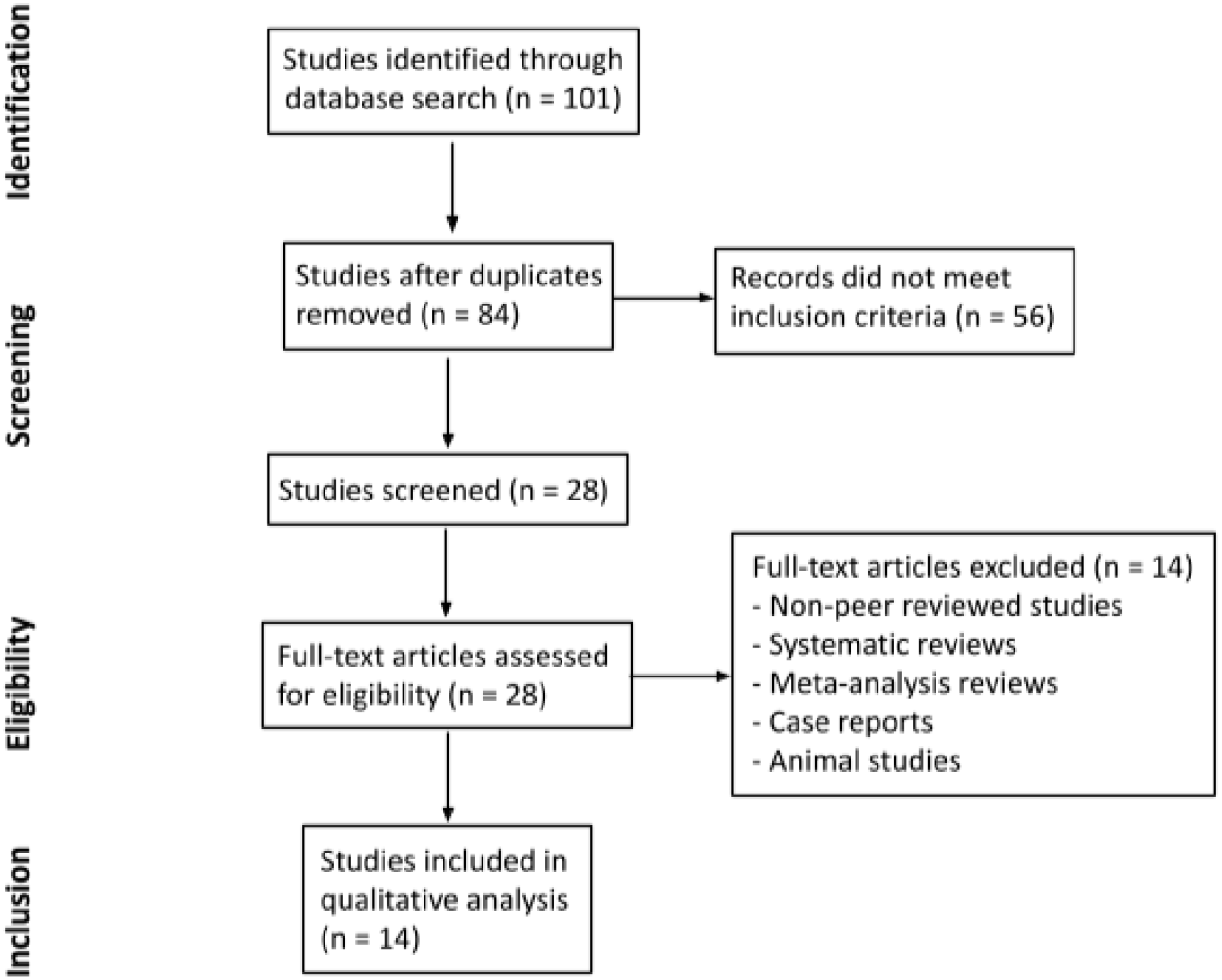
Flow chart of the study selection and inclusion process.

Participant ages ranged from 12 to 68 years. Follow-up periods ranged from 3 hours to 5 years, with four studies having follow-up of less than one week (three hours to five days), eight studies with one to six months, and two studies from three to five years.

All studies used intravenous ketamine infusions as the intervention. Five of the 14 studies used escalating, titrated doses, while the other nine studies used the same dosage throughout the duration of the intervention. Duration of analgesia ranged from five minutes to ten days based on the dosage of ketamine used and time length measured in each study. Most studies tailored ketamine dosages based on individual weight, with doses ranging from 0.15 to 7 mg/kg/hour.

Kirkpatrick *et al*.^24^, Kiefer *et al*.^25^, and Goldberg *et al*.^26^ did not calculate dosages based on patient weight, and instead used dosages between 10 to 200 mg/hour.

All studies but one used some form of objective pain scale such as the visual analog scale (VAS) or verbal numeric rating scale (NRS), which range from 0 (no pain) to 10 (worst possible pain). Instead of a pain score, Kirkpatrick *et al*.^24^ measured pain threshold, which is defined as the amount of stimulation before the sensation of pain is experienced. In 13 of the studies, ketamine infusion resulted in a decrease in pain scores or threshold. Kiefer *et al*.^25^ did not find a significant change in pain relief in their sample of patients with long-standing, severe CRPS. They posit this may be due to non-NMDAR-mediated mechanisms that exist in patients with severe CRPS refractory to numerous treatments essentially rendering ketamine non-effective.

Other measured outcomes included morphine-equivalent intake, plasma levels of ketamine, quality of life, functional improvement, cognitive effects, and side effects of ketamine infusion. Sheehy *et al*.^27^ measured oral morphine-equivalent opioid intake, and found that while ketamine reduced pain intensity, it did not reduce overall opioid intake. Goldberg *et al*.^26^ also measured plasma levels of ketamine, and found that pain relief correlated with maximum plasma levels of ketamine and norketamine, indicating that downstream metabolites may play a role in therapeutic effect. Kiefer *et al*.^28^ and Schwartzman *et al*.^29^ also measured quality of life (QOL) as an outcome. Data from Kiefer *et al*.^28^ supported a significant improvement in QOL in the majority of patients at 3- and 6-months following treatment, while Schwartzman *et al*.^29^ found that there were no significant changes in QOL scores following treatment in either the treatment or placebo groups.

Functional improvement, including ability to work, activity level, active range of motion, and effect on movement disorder, was measured in four studies. Kiefer *et al*.^28^ showed that ability to work was significantly improved 3 months post-treatment, with an even greater improvement by 6 months. Schwartzman *et al*.^29^ found that based on activity watch data, there were no significant changes in activity level between pre- and post-treatment, but the number of nighttime awakenings in the ketamine group decreased by 85% post-treatment. Puchalski *et al*.^30^ and Sigtermans *et al*.^31^ did not show that treatment caused functional improvement in active range of motion and ability to walk without support.

Cognitive effects were assessed in Koffler *et al.*,^32^ which found that there were significant improvements in brief auditory attention and processing speed, no changes in learning, memory, nor motor speed, and a slight decline in motor strength. Side effects of ketamine infusion were directly assessed in 7 of the 14 studies. The majority of studies reported mild general symptoms of fatigue, headache, and nausea. Several studies also reported participants with side effects of vomiting, feeling of inebriation, and sedation. Two studies by Correll *et al*.^33^ and Sigtermans *et al*.^31^, reported patients with mild-to-moderate psychomimetic effects, such as hallucinations.

The findings from all 14 included studies are summarized in Table 1. Overall, a majority of the fourteen studies produced positive results for the effect of ketamine infusions on improved pain control and function for the treatment of CRPS. This was achieved by improved VAS or NRS pain scores or pain threshold. Increased quality of life and positive changes in physical and cognitive function in CRPS patients treated with ketamine was also reported. One study did not show a significant change in pain relief with ketamine infusion in patients with severe disease.

**TABLE 1.** Summary of findings from included studies.

| Author | Year | Study | Sample Size | Average Age (Range) (years) | Follow-up Period | Intervention | Ketamine Dose Used | Measures | Conclusions |
| --- | --- | --- | --- | --- | --- | --- | --- | --- | --- |
| Sheehy *et al.*<sup>27</sup> | 2015 | Longitudinal, cohort | 63 | 15 (12-17) | 1 month | Subanesthetic ketamine infusions - 4–8 h per day for 3 days | 0.1–0.3 mg/kg/h | Self-reported pain scores, morphine-equivalent intake, side effects | Ketamine significantly reduced pain intensity and yielded greater pain reduction, but did not change overall morphine-equivalent intake. Ketamine was not associated with psychotropic effects at doses given. |
| Schwartzman *et al.*<sup>29</sup> | 2009 | Randomized control trial | 19 | 42 (24-60) | 3 months | IV Ketamine infusions 4h (25ml/h) daily for 10 days | 0.35 mg/kg/h | Pain questionnaires, activity watch data, quantitative sensory testing, quality of life (QOL), side effects | IV ketamine administered in an outpatient setting resulted in significant reductions in many pain parameters, while the placebo group demonstrated no treatment effect in any parameter. These results warrant a larger RCT using higher doses of ketamine and a longer follow-up period. |
| Sigtermans *et al.*<sup>31</sup> | 2009 | Randomized control trial | 48 females | 45 (33-57) | 3 months | Individually tailored IV low dose ketamine infusions held for 4.2 days | 22.2 ± 2.0 mg/h/70kg (maxed at .43mg/kg/hr ) | Pain score, skills questionnaire, AROM, touch sensitivity, skin temperature, volumetric measurement, LFTs, side effects | In a population of mostly chronic CRPS-1 patients with severe pain at baseline, a multiple-day ketamine infusion resulted in significant pain relief without functional improvement. Treatment with ketamine was safe with psychomimetic side effects that were acceptable to most patients. |
| Puchalski *et al.*<sup>30</sup> | 2016 | Observational study | 5 females | 34 (25-45) | 1.5 months | 4 Hour Infusion of Anesthetic IV ketamine dosage over 10 days | 1.5 mg/kg/h | Pain score, presence of allodynia, ROM, ability to walk without support | This beneficial analgesic effect was confined to 1.5-2.5 months after treatment and then pain relapsed to the baseline level. These results show a short-term analgesic effect for this therapy, with no effect on movement and function of the affected limbs. |
| Sigtermans *et al.*<sup>34</sup> | 2010 | Observational study | 10 | 43.2 ± 13.0 | 3 hours | 7 IV 5 minute long low-dose ketamine infusions with increasing doses at 20 min intervals | .02mg/kg - .15mg/kg | Spontaneous pain ratings, VAS response to experimental heat stimuli | While ketamine's effect on acute experimental pain is driven by pharmacokinetics, its effect on CRPS pain persisted beyond the infusion period when drug concentrations were below the analgesia threshold for acute pain. This indicates a disease modulatory role for ketamine in CRPS-1 pain, possibly via desensitization of NMDAR in the spinal cord or restoration of inhibitory sensory control in the brain. |
| Correll *et al.*<sup>33</sup> | 2004 | Retrospective case notes analysis | 33 | 40 (20-68) | 3 years | Continuous subanesthetic IV Ketamine infusion for 4.7 days | 23.4mg/kg | Pain scale, degree of relief, side effects | Limited subanesthetic inpatient infusions of ketamine may offer a promising therapeutic option in the treatment of appropriately selected patients with intractable CRPS. Further studies are needed to establish the safety and efficacy of this novel approach. |
| Kirkpatrick *et al.*<sup>24</sup> | 2020 | Observational study | 114 (89 females and 25 males) | 39 (9-50) | 3 days | Escalating dose ketamine IV infusion | Start at 60 mg/hr, titrated up to 200 mg/hr on Day 4 | Pain threshold | Four days of treatment are sufficient for the treatment of CRPS of the lower extremities. For the upper extremities, >4 days may be required. This was the first study to utilize quantitative sensory testing to direct the treatment of a chronic pain disorder. |
| Dahan *et al.*<sup>35</sup> | 2010 | Randomized control trial | 60 | 46 ± 12 | 2.5 months | 100-h titrated ketamine or placebo infusion | .43mg/kg/hr | Pain scores, blood samples | Long-term S(+)-ketamine treatment is effective in causing pain relief in CRPS-1 patients with analgesia outlasting the treatment period by 50 days. These data suggest that ketamine initiated a cascade of events, including desensitization of excitatory receptor systems in the central nervous system, which persisted but slowly abated when ketamine molecules were no longer present. |
| Patil *et al.*<sup>36</sup> | 2012 | Retrospective case notes analysis | 49 | 45 (18-68) | 5 years | IV ketamine infusions, pretreatment with midazolam and ondansetron | 0.5 mg/kg given over 30–45 minutes | Pain score, long-term pain relief, previous interventions, side effects | In patients with severe refractory pain of multiple etiologies, subanesthetic ketamine infusions may improve VAS scores. In half the patients, relief lasted for up to 3 weeks with minimal side effects. |
| Kiefer *et al.*<sup>28</sup> | 2008 | Open label trial | 20 | 30.4 (14-48) | 6 months | 5-day anesthetic ketamine dosage | 7 mg/kg/h | Pain score, effect on movement disorder, QOL, ability to work | Anesthetic ketamine in previously refractory CRPS patients lead to benefits in pain reduction, associated CRPS symptoms, improved quality of life, and ability to work. A RCT will be necessary to prove its efficacy. |
| Goebel *et al.*<sup>37</sup> | 2015 | Observational study | 5 | 44 (37-55) | 6 months | Low-dose, 4.5-day RK infusion | .9mg/kg/hr | Pain score, side effects, LFT/CBC | IV infusion of subanaesthetic doses of S+K over 4.5 days can substantially reduce pain in long-standing CRPS for several weeks. |
| Koffler *et al.*<sup>32</sup> | 2007 | Observational study | 9 | 29 (19-41) | 1.5 months | 5 days of anesthetic ketamine infusion | 3–7 mg/kg/h | Acute and overall pain, brief attention and processing speed, cognitive domains, motor strength | Deep ketamine therapy is effective for relief of pain CRPS I. There were no adverse cognitive effects of extended treatment with deep ketamine infusion. No definitive conclusions could be drawn about the relationship between mood and personality factors and the presence of CRPS I. |
| Kiefer et al.<sup>25</sup> | 2008 | Clinical trial | 4 | 25<br>(18-43) | 2 days | S(+)-ketamine<br>-infusions,<br>titrated over<br>a 10-day<br>period | 50<br>mg/day-500<br>mg/day | VAS pain,<br>thermo- and<br>mechanical<br>detection<br>thresholds,<br>side effects | S(+)-ketamine can be gradually titrated to large doses (500 mg/day) without clinically relevant side effects. There was no pain relief or change in QST measurements in this series of long-standing severe CRPS patients. |
| Goldberg et al.<sup>26</sup> | 2010 | Clinical trial | 16 | 33<br>(17-47) | 5 days | Titrated<br>ketamine<br>infusion<br>maintained<br>for 5 days | 10-40<br>mg/hour | Pain scale,<br>plasma levels<br>of ketamine | The pain relief experienced on Day 2 of the infusion continued to improve over the 5-day infusion period and correlated with the maximum plasma levels of ketamine and norketamine. Downstream metabolites of ketamine and norketamine might be playing a role in its therapeutic efficacy. |

## Discussion

A double-blind placebo-controlled study observing the effects of outpatient IV ketamine for the treatment of CRPS was conducted by Schwartzman *et al*.^29^ In the study, all subjects diagnosed with CRPS with pain for more than 6 months and who failed conservative therapy such as nerve blocks, opioid analgesics, and NSAIDs were included in the trial. Subjects receiving ketamine treatment were infused with 25 ml/h daily for 4 hours for a 10-day treatment period. In this study, all subjects that received both ketamine and the placebo were given treatment with clonidine to suppress any side effects. Results showed that the IV ketamine group had a statistically significant reduction in their pain parameters while the placebo group showed no treatment effect in the pain parameters. To evaluate pain, all subjects completed two pain questionnaires prior to treatment and one pain questionnaire every week for 12 weeks post-treatment. For this study, the NRS pain questionnaire was used which was scaled from 0-10. For the ketamine group, significant decreases in all parameters post-treatment were observed. Of note, the decreases in pain parameters lasted the entirety of the 12-week treatment course. Pain in the affected area, burning pain, pain when touched or brushed lightly, and overall pain level were all parameters that had significant decreases in the ketamine group. Given the decrease in pain scores in these specific parameters, the use of ketamine likely had significant overall improvement in quality of life and may be considered as an option for patients who have failed conservative measures such as the subjects in this trial.

Another study by Sigtermans *et al*.,^31^ explored the use of ketamine on pain relief for patients with CRPS-1. In the study, 60 CRPS-1 patients were included in a double-blind randomized placebo-controlled trial. It is notable that the included group in this study consisted of patients with a diagnosis of CRPS-1 referred to the clinic regardless of previous history of response to conservative treatment. In the other studies discussed in this review, many patients included in a ketamine trial had failed conservative treatment for a minimum number of months prior to receiving ketamine infusion. Patients were given a 4-day infusion of ketamine with a dose of 22 mg/h/70kg. All subjects were evaluated based on pain scores over the 12-week study period.

Results showed patients receiving ketamine had significantly lower pain scores than patients receiving placebo treatment. Although significance was established in the ketamine group, it is notable that by week 12, the significant inter-group difference in pain relief was lost. Patients had no functional improvement with the ketamine treatment and also experienced mild to moderate psychomimetic side effects.

A longitudinal study by Sheehy *et al*.^27^ observing the effects of sub anesthetic ketamine infusions for the treatment of children and adolescents with chronic pain was also reviewed. In this study, the effects of ketamine on both CRPS and other chronic pain syndromes were compared. 63 children received IV administration of subanesthetic doses of ketamine in an outpatient setting with dosing ranging from 0.1 to 0.3 mg/kg/h that lasted for 4-8 hours per day. Similar to the other studies, the primary outcome measure was change in pain scores using a numeric rating scale. A primary result of the study was that the effect of ketamine on pain scores varied based on the subject’s pain diagnosis. Overall, the use of ketamine resulted in greatest pain score reductions in patients with CRPS than any other chronic pain condition. In addition, multivariate analysis was able to further qualify that CRPS was a significant predictor of higher pain score reductions after ketamine treatment. Although targeting pediatric patients, this study further provides validity that ketamine may be safely used for chronic pain syndromes, particularly CRPS.

Prolonged painful stimulus causes an increased release of glutamate from nociceptive afferents onto the dorsal horn neurons. This glutamate stimulates NMDA receptors on second-order neurons which lead to increase in pain intensity (windup) and cause central sensitization. Corelle *et al*. posit that blocking these NMDA receptors is what propagates ketamine’s ability to treat CRPS. Temporally, there are results showing a patient with 20 years of CRPS having full suppression of pain after infusion of ketamine, pointing towards a neuromodulatory effect. Mean survival of pain relief was found to be 3 to 6 months with only one dose and up to one to three years with a second infusion.^33^ Interestingly, the ideal infusion rate was found to be 15-20 to 20-25mg/hr over the course of 10 to 20 days, with those receiving more than 35mg/hr categorized as non-responders or had exacerbated side effects. Side effects are a primary concern with ketamine therapy with potential neurotoxic effects observed in animal studies. It is yet to be seen how well these findings correlate in humans, therefore the predominant approach is to dose at constant low rates. The most common side effects reported was feeling intoxicated followed by hallucinations as seen in 18% of the study population. Although the results are highly encouraging, the study population contained a large number of early CRPS diagnosis with no established protocol for treatment and assessment of results.

At the pharmacological level, Sigtermans *et al*.^34^ studied the S(+) variant of the compound due to its greater availability and two fold greater analgesic potency than S(-) or racemic mixtures.

They found a plasma concentration driven effect on acute pain which showed an “on-off” phenomenon that inhibits NMDR in the central pain circuit. The study tested 10 females and administered seven IV infusions that were 5 minutes long of low-dose ketamine with increasing doses at 20 min intervals. Of note, the study found it difficult to blindly administer S(+) ketamine and compare it to placebo due to two main reasons. One, the effects of ketamine on patient affect are apparent to the administrator. Second, creating a placebo with similar behavioral effects (i.e., drowsiness) is difficult due to mixing of drugs that might have independent pain modification processes. It is advocated that a de-chronification of pain occurs secondary to a long-term desensitization of upregulated NMDA receptors. Further research is to be done to differentiate this mechanism of action versus a top-down inhibitory control of pain sensory systems.

Escalating doses of ketamine infusion, starting at 60mg/hr to 200mg/hr, based on quantitative sensory testing to direct treatment, demonstrate that ketamine has its most potent effect on those areas of the body that are most affected by pain due to CRPS. A distinction is made when treating extremity pain in patients with CRPS, with pain threshold plateaus being realized between 3 and 4 days, and greater than 4 days of ketamine treatment for patients with CRPS of the lower and upper extremities, respectively.^24^ Similarly, long-term (100-h) escalating doses of ketamine infusion starting at 5mg/hr/70kg to 20mg/hr/70kg demonstrate effectiveness in pain relief for CRPS-1 patients, with analgesia outlasting the treatment period by 50 days.^35^ This observation suggests that ketamine initiates a cascade of events, including desensitization of excitatory receptor systems in the CNS which persist but abate slowly, even when ketamine molecules are no longer detectable in the body.

A 5-year retrospective analysis studying the efficacy of outpatient ketamine infusions revealed that 50% of patients with severe refractory pain of multiple etiologies realized pain relief for up to 3 weeks with minimal side effects after receiving mean ketamine doses per infusion of 0.9 (±0.4) mg/kg with a median duration between infusions of 233 days. All patients were pretreated with midazolam, ondansetron, and an initial ketamine dose of 0.5mg/kg given over 30 to 45 minutes. If the initial dose was effective, it was continued in subsequent infusions. They were then scheduled to receive treatment dosages every 3-4 weeks per pain clinic protocols.^36^ If well-tolerated, the dose was increased at subsequent infusions to the highest tolerated dose producing analgesia without unacceptable side effects. While these patients’ long-term results are unavailable per this protocol, Kiefer *et al*.^28^ noted long-term complete pain relief, 6 months post-treatment, in 50% of patients with advanced and refractory CRPS who underwent a 5-day anesthetic ketamine infusion protocol. Anesthesia was induced by bolus injection of ketamine (1–1.5 mg/kg) and midazolam (2.5–7.5 mg). Tracheal intubation was facilitated by vecuronium (0.1 mg/kg). Treatment was maintained by infusions of ketamine over 5 days, starting at 3 mg/kg/h, followed by gradual daily titration up to a final dose of 7mg/kg/h. Midazolam was co-administered and adjusted as required to obtain a stable level of deep sedation and to attenuate ketamine-specific agitation.

Ketamine infusions can provide significant analgesia to patients with CRPS that is refractory to conservative methods of pain management. One observational study by Goebel *et al*. evaluated the short- and long-term analgesic effects of 4.5-day, low-dose (0.9mg/kg/hr) racemic ketamine (RK) infusions in five patients with refractory CRPS.^37^ The study authors found that intravenous infusion of low, subanesthetic doses of RK over a 4.5-day period can substantially reduce pain in patients with CRPS with 60% of patients reporting substantial pain relief during the treatment period.^37^ The median pain intensity at the beginning of the treatment period was NRS 8.5, and was reduced to NRS 5 on the last treatment day.^37^ The authors concluded that low-dose RK infusion over the duration of 4.5 days provided beneficial analgesia for short durations, but further prospective trials are needed to understand long-term pain control with multiple repeat ketamine infusions.^37^ Koffler *et al*. conducted an observational study to understand pain control and neurocognitive effects of a 5-day anesthetic ketamine infusion in patients with CRPS.^32^ The study evaluated intellectual and academic abilities, executive functioning and processing speed, attention, learning, memory, and motor functioning in nine patients pre-treatment and 6-weeks post-treatment. The study population demonstrated significant pain reduction (present pain index (PPI); t(8) = 2.393, p = .044) without adverse cognitive effects of ketamine infusion at 6-weeks post-treatment.^32^

Ketamine, norketamine, and the downstream metabolites likely play an important role in the analgesic properties and efficacy of ketamine infusions. Goldberg *et al*.^26^ conducted an observational study to determine the effects of a 5-day moderate dose, continuous racemic ketamine infusion while evaluating for S-ketamine, R-ketamine, S-norketamine, and R-norketamine plasma levels.^26^ Sixteen patients with CRPS enrolled in an observational cohort study were assessed daily for pain and plasma concentrations of ketamine and norketamine. Concentrations were measured before initiation of infusion, at several time points during the infusion, on Days 2 through 5 of the infusion, and 60 minutes after the conclusion of infusion on Day 5. The study authors found that the ketamine infusion resulted in significant pain reduction by the second day of infusion which correlated with peak plasma concentrations, supporting the antinociceptive properties of the drug.^26^

While several studies have shown ketamine infusions to be an effective analgesic for pain management in patients with refractory CRPS, a pilot open-label study by Kiefer *et al*.,^25^ analyzing the efficacy of subanesthetic ketamine infusion in refractory CRPS patients found that treatment did not offer pain relief. Four study patients with refractory CRPS were enrolled to receive continuous S+-ketamine infusions gradually titrated (50mg/day - 500mg/day) over a 10-day period. Pain intensity and side effects on 100-mm visual analog scale (VAS) during a day baseline, over 10 treatment days, and 2 days post treatment were measured. The subjective pain relief with ketamine infusions was 4.6 ± 1.6mm on the 100-mm VAS.^25^ Over the 10 treatment days, there was no significant difference in pain relief between the treatment and baseline pain scores.

### Future Studies

Overall, the included studies generally reported adequate pain relief with varying protocols of ketamine infusion for treatment-resistant CRPS. Heterogeneity was significant in these studies due to variations in dosage, protocol, number of subjects, combinations of pharmacological agents administered and rescue analgesics. Despite these differences, some valuable trends can be observed and may provide direction on future studies for the utility of ketamine infusion. Of interest would be identifying the ideal time course in treatment regimen to use ketamine for maximal efficacy. Also, dosage stratification should be explored based on age, severity of pain as per numerical scale ratings and time of living with disease. With new innovations in pain management such as neurostimulation and implantable pumps, a comparative study with ketamine should also be undertaken to understand the safest way to address pain control.

Ketamine’s concurrent influence on psychology should be further understood as pain is a complex entity that affects numerous functionalities.

### Strengths and Limitations

Strengths of this review included select studies that were published in peer-reviewed journals with thorough methodology and low dropout rates. However, this systematic review also had several notable limitations, including significant heterogeneity of the included studies in regard to study design, sample size, dosing strategies, and tools used to measure variables and outcomes. Among the 14 studies reviewed, yielding a total of 455 included patients, several confounding factors were identified including variable ketamine dosing strategies and durations of treatment, observational design in studies, and use of other nonopioid multimodal analgesia. Study subjects were assessed for pain scores and other outcomes at varying intervals, which may influence outcomes of recall, in turn, increasing the risk of recall bias. Moreover, pain is subjective and challenging to measure despite utilization of validated scales.

## Conclusion

Patients who received ketamine infusion for treatment-resistant CRPS reported adequate pain relief with treatment. Given the results, this suggests that ketamine infusion may be a useful form of treatment for patients with no significant pain relief with other conservative measures. Although pain scores were reduced with ketamine use in all but one study reviewed, parameters such as functional improvement and quality of life were not as well studied. In addition, one of the included studies observed the effects of ketamine in pediatric patients with chronic pain syndromes signaling a scope for use in younger patients. Future large-scale studies, such as randomized double-blind placebo-controlled trials, must be conducted to better correlate the use of ketamine infusion in CRPS patients with improved pain scores, changes in parameters such as quality of life and activities of daily living as well as effectiveness by age cohorts.

## Data Availability

All articles accessible as cited in the manuscript through their respective citations.

